# Analysis of the intensity of the COVID-19 epidemic in Berlin towards an universal prognostic relationship

**DOI:** 10.1101/2021.01.03.21249117

**Authors:** D. Below, J. Mairanowski, F. Mairanowski

## Abstract

The present work is a continuation and development of research on prediction and analysis of the spread of the COVID-19 epidemic.

The proposed model adequately describes the development of the coronavirus epidemic with insufficient adherence to quarantine and social distancing. The transition from the absolute number of infected persons to their relative number per inhabitant of a settlement makes it possible to obtain universal calculation ratios.

In performing the calculations, the choice of the date of the beginning of the epidemic is of great importance. Recommendations are given on how to determine the date of the beginning of the epidemic based on the analysis of statistical data on the spread of the epidemic. The coefficient of virus transmission rate k included in the calculated prognostic relation depends on the population size and the type of virus strain in the settlement in question. A simple ratio for calculating this coefficient as a function of population size is proposed.

Control calculations performed using only a single empirical coefficient showed high accuracy. The calculated curves for Germany, Berlin, and its districts agree well with the corresponding statistical data. The correlation coefficients between the corresponding curves reach values of 0.93 to 0.97. The further development of the model should thus go in the direction of identifying causal links between the intensity of the epidemic and the main factors affecting this process. Some of these factors are related to the characteristics of the population’s behaviour and the infrastructure of cities. The increase in the incidence in areas with a large percentage of the population rooted in Islamic countries is one of the main factors determining the development of the epidemic in Berlin. In order to explain and clarify this conclusion, it is necessary to make further assumptions about the possible emergence of a new strain of coronavirus in Berlin and in Germany and, accordingly, about the possibility of new epidemic waves. A preliminary ratio for predicting the spread of the epidemic under conditions of simultaneous existence of both strains of coronavirus is given.

Simplicity of the proposed prognostic method and high accuracy of the results allow to recommend it as an effective tool for operative analysis of various measures aimed to control the spread of COVID-19 epidemic including mass vaccination of population.

## Formula development and resulting analysis

The present work is a continuation and development of research on predicting and analysing the spread of the COVID-19 epidemic [1], [2]. In the previous work it was found that the intensity of virus spread can be estimated using analytical dependence obtained from the solution of a simplified system of differential equations describing the regularities of epidemic development under quarantine. The obtained analytical dependence of the change in the number of infected persons on time is written in the form [1]:

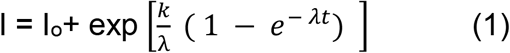

In which:

Io - the number of infected patients at the beginning of the calculation period,

t - time in days, counted from the beginning of the calculation period

k - coefficient characterizing the transmission rate of the virus, which depends on both the nature of the virus and the size of the settlement for which the prediction is made,

λ *-* intensity factor of decrease in contacts of infected patients with with persons who potentially can get infected by means of quarantine and other preventive measures.

As a result of differentiating (1) by time and equating the first derivative to zero, we obtained a formula for determining the time of maximum growth of infected persons, counting from the beginning of the epidemic

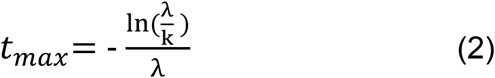

and the value of the maximum daily increase in the number of persons affected by the virus

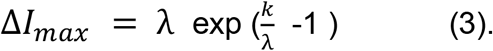

The question of choosing a date for the beginning of the epidemic deserves separate consideration. In the previous work, August 14^th^ was taken as this date for Berlin, the date of active return from vacations of a large number of holidaymakers and the beginning of classes in schools. However, it is more rational to choose the date of the beginning of the calculation period from the analysis of real data on the intensity of the daily increase in the number of infected persons. Figure 1 shows a graph of the daily change in the number of infections in Berlin for the period August 28^th^ to September 24^th^, 2020 [3].

**Fig.1:**
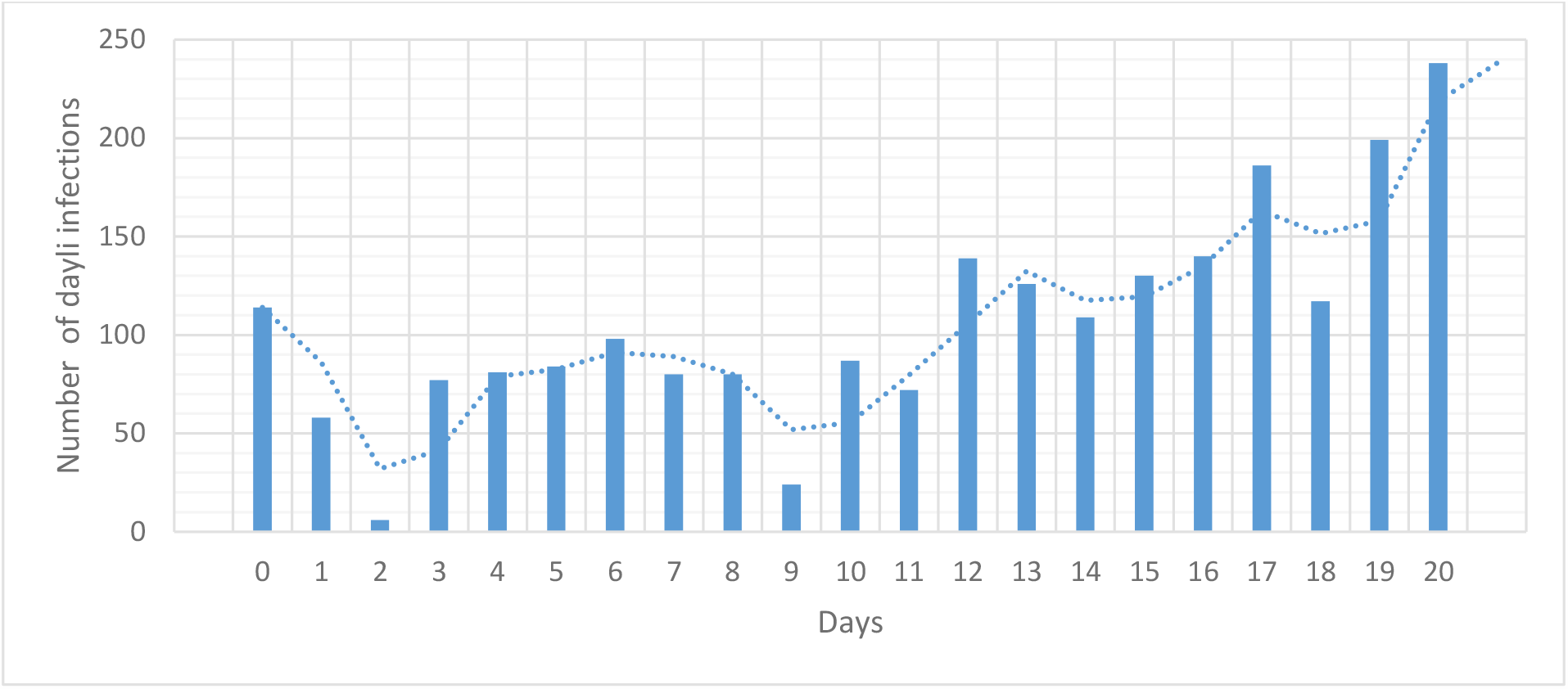
Number of infected persons at the beginning of the epidemic

From the analysis of this graph it was accepted that the steady tendency of the increase is observed from day 17, i.e. from 18.09.2020. Considering that the disease begins to manifest itself noticeably in about 14 days after infection, we chose 04.09.2020 as the beginning of the second wave of the epidemic. The values of the model coefficients k and λ were not different from those used in the previous study, that is k = 0.4 1/day, λ = 0.035 1/day. Figure 2 shows the comparison of the results of calculations according to dependence (1) with the observational data from the beginning of the calculation period to the date of this study (18.12.2020) [3].

**Fig.2:**
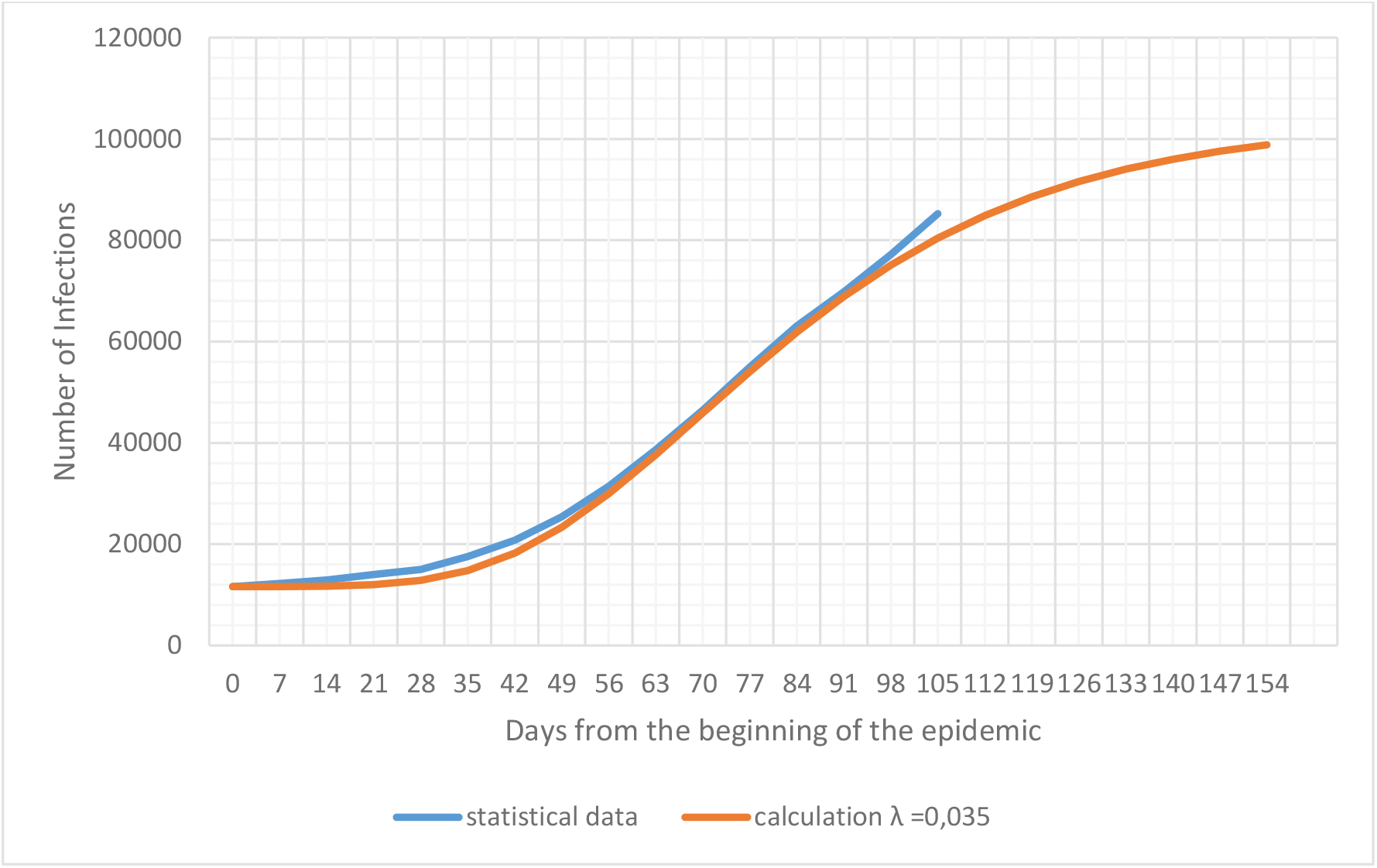
Intensity of the epidemic in Berlin

The correlation coefficient between both curves is r = 0.96. At the same time, after about 84 days from the beginning of this epidemic, i.e. from November 27^th^, one can notice a rather significant difference between the real data and the calculated ones. Moreover, starting from this period, the increase in infections becomes more intense than in previous periods. Consequently, there appeared a new factor which influences the intensity of virus transmission. Taking into account that in this time period there were no significant changes related to preventive quarantine measures, we can draw a preliminary conclusion about a possible mutation of the virus, which led to the intensification of the epidemic. This assumption is indirectly confirmed by data on a significant increase in the spread of the virus in Great Britain as a result of its mutation [4]. However, virologists in Germany have not detected the emergence of a new strain of the virus in the country (at the time of writing this paper). Analysis of these data shows a very important pattern discovered at the beginning of the first epidemic wave by studying the spread of the epidemic in New York [2]. The spread of the virus is much easier to prevent at a very early stage of its emergence. Once the level of infection of a population reaches a certain scale, it becomes very difficult to reduce the intensity of the spread of the virus. An additional difficulty in preventing the spread of the epidemic is related to the fact that the virus often does not become active until 10 to 14 days after infection.

The number of infected persons depends on the size of the population of a country, city or individual area. In order to be able to compare the intensity of the epidemic among themselves, it is advisable to analyse the specific number of infections per inhabitant, that is, the frequency of infection, for example in percent. Figure 3 shows statistical data on the specific intensity of the epidemic in percent for Berlin as a whole and for some of its districts. With this comparison the practical coincidence of the curves for Berlin and Charlottenburg-Wilmersdorf is immediately striking. As for the districts of Neukölln and Mitte, the intensity of spread in them is significantly higher than in Berlin, and for the district of Treptow-Köpenick, on the contrary, significantly lower.

**Fig.3:**
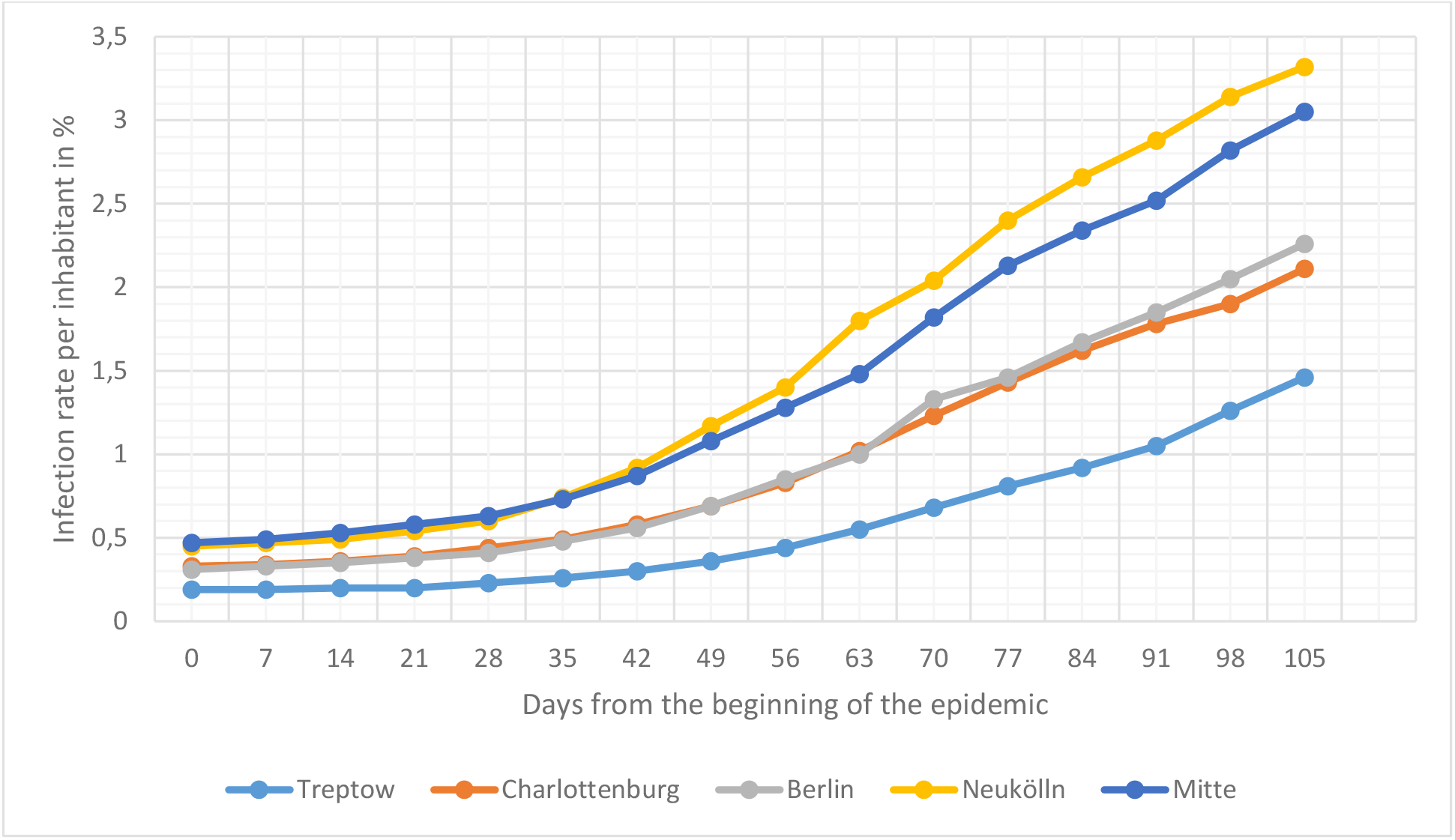
Statistical data on the spread of the epidemic in Berlin and some of its areas

**Fig.4:**
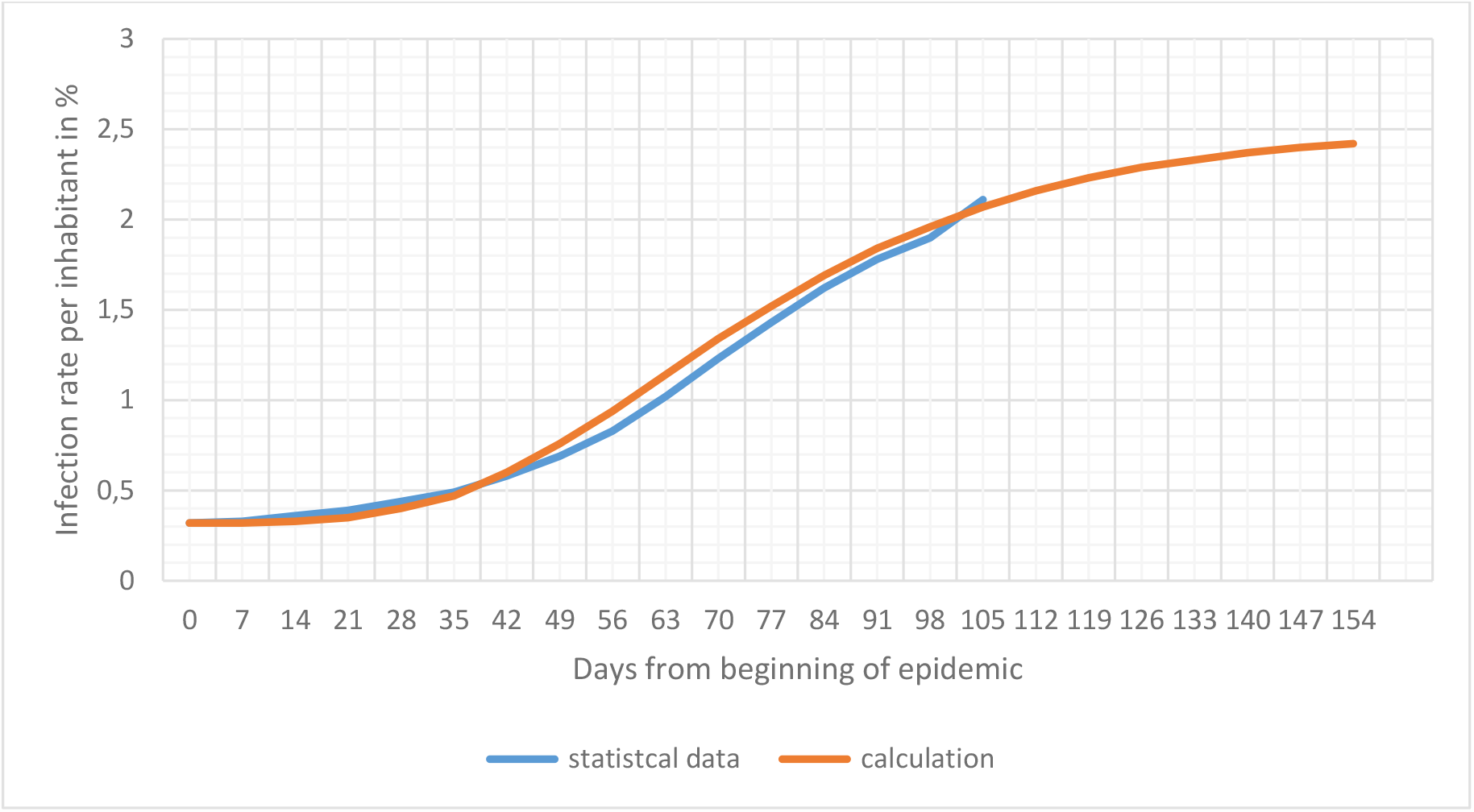
Relative growth of the epidemic in the Charlottenburg-Wilmersdorf district

**Fig.5.**
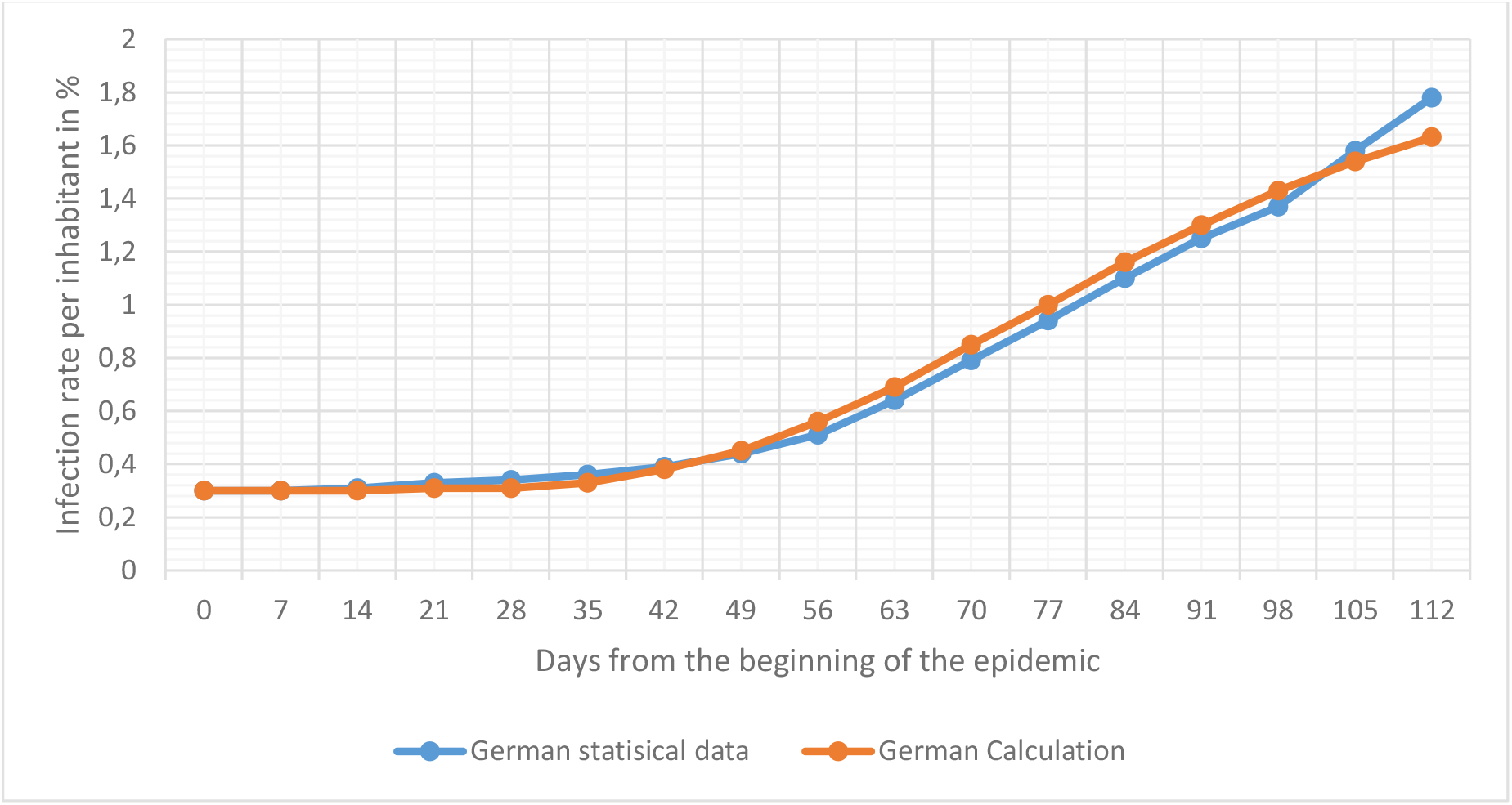
Relative growth of the coronavirus epidemic for Germany

These differences, as pointed out in our previous work [1], are associated with certain characteristic features of the social structures of different regions. So, in particular, from the analysis of statistical data was obtained that dependence of the number of infected people for each of the 12 districts in Berlin on the relative size of the population from the member countries of the Organization of the Islamic Conference (OIC) (Turkey, Arab States, some African countries). Statistics on the number of infections were taken for 1^st^ November 2020 according to data on the demographic composition of the inhabitants of each district according to [5]. The high correlation coefficient (r = 0.93) shows that the connection between these parameters is not only statistically reliable, but also practically functional. For Berlin, the relative number of foreigners from OIC countries was 12%, for the Charlottenburg-Wilmersdorf district it was 11.5% (at the end of 2019).

The main conclusion that follows from the analysis of Fig. 3 is extremely important and is that the spread of the virus is not determined by random characteristics, but is quite deterministic. In order to develop a reliable method for predicting the development of an epidemic, it is necessary to establish the main factors determining the intensity of this process.

First of all, however, it is necessary to transform the main estimated dependence so that it takes into account the real population size. In doing so, we will proceed from the confirmed assumption that, for the city of Berlin with a population of 3.57 million, the transmission rate coefficient for the strain in question is 0.4 1/day. The coefficient λ does not depend on the population size and is determined, as already mentioned, only by the quarantine conditions.

Taking these assumptions and using equation (1), we can obtain a simple ratio to calculate the coefficient k depending on the number of inhabitants:

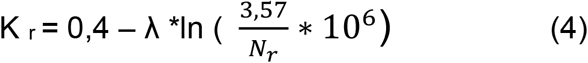

in which k_r_ is the calculated coefficient for the point with population N_r_.

λ = 0,035 1/day

Thus, we find the ratio for calculating the number of infected persons per one inhabitant of the settlement as a percentage:

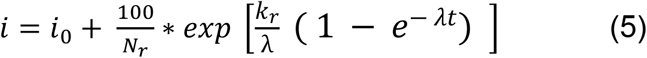

Using relation (5), calculations were made for Germany, Berlin and all of its 12 districts. The results of the calculations were compared with the corresponding statistical data on the spread of the virus. The initial values of *i*_0_ were also taken according to the observation data Fig.4 shows a graph of the relative number of infected persons for the Charlottenburg-Wilmersdorf district.

The coherence of the calculated and statistical data is quite good. The correlation coefficient between both curves is r = 0.967. As already noted, this graph practically coincides with the similar graph for Berlin (except for the data for December 4^th^ and December 11^th^, which has already been pointed out when discussing the graph in Fig. 2).

Comparison of the results of calculations according to the ratio (5) and statistical data for Germany [6]. The analysis of initial statistical information showed that in Germany the new virus epidemic started about a week earlier than in Berlin. Therefore, the date of August 28^th^, 2020 was taken as the beginning of the epidemic in calculations for Germany.

The correlation coefficient between statistical data and calculation results for Germany is r = 0.937. This graph, as well as the previous ones, shows that starting from the end of November there is also a tendency of more intensive growth of the epidemic. In general, the intensity of the epidemic development for Germany is somewhat weaker than for Berlin. The presented data of epidemic growth per inhabitant in Germany practically coincide with similar data for Berlin Reinickendorf district.

One of those districts of Berlin where the development of the epidemic is the least intense is Treptow-Köpenick. Fig. 6 shows a comparison of the results of the calculation according to dependence (5) with statistical data. The epidemic in this area of Berlin has several peculiarities compared to other areas. First, the beginning of the epidemic was observed about a week later than in Berlin. In addition, about 70 days after the beginning of the epidemic, the intensity of the epidemic continued to increase, and this trend intensified at the beginning of December (i.e. after three months from the beginning of the second wave of the epidemic). As a consequence, the calculated maximum values of the number of infected persons are significantly lower than the real statistical data. In spite of a sufficiently high correlation coefficient between the calculated and statistical data (r = 0,935), we should admit that some correction of the calculated dependence is necessary for this area. It is possible that such intensive growth of the epidemic is associated with the emergence of a new strain of the virus.

**Fig.6:**
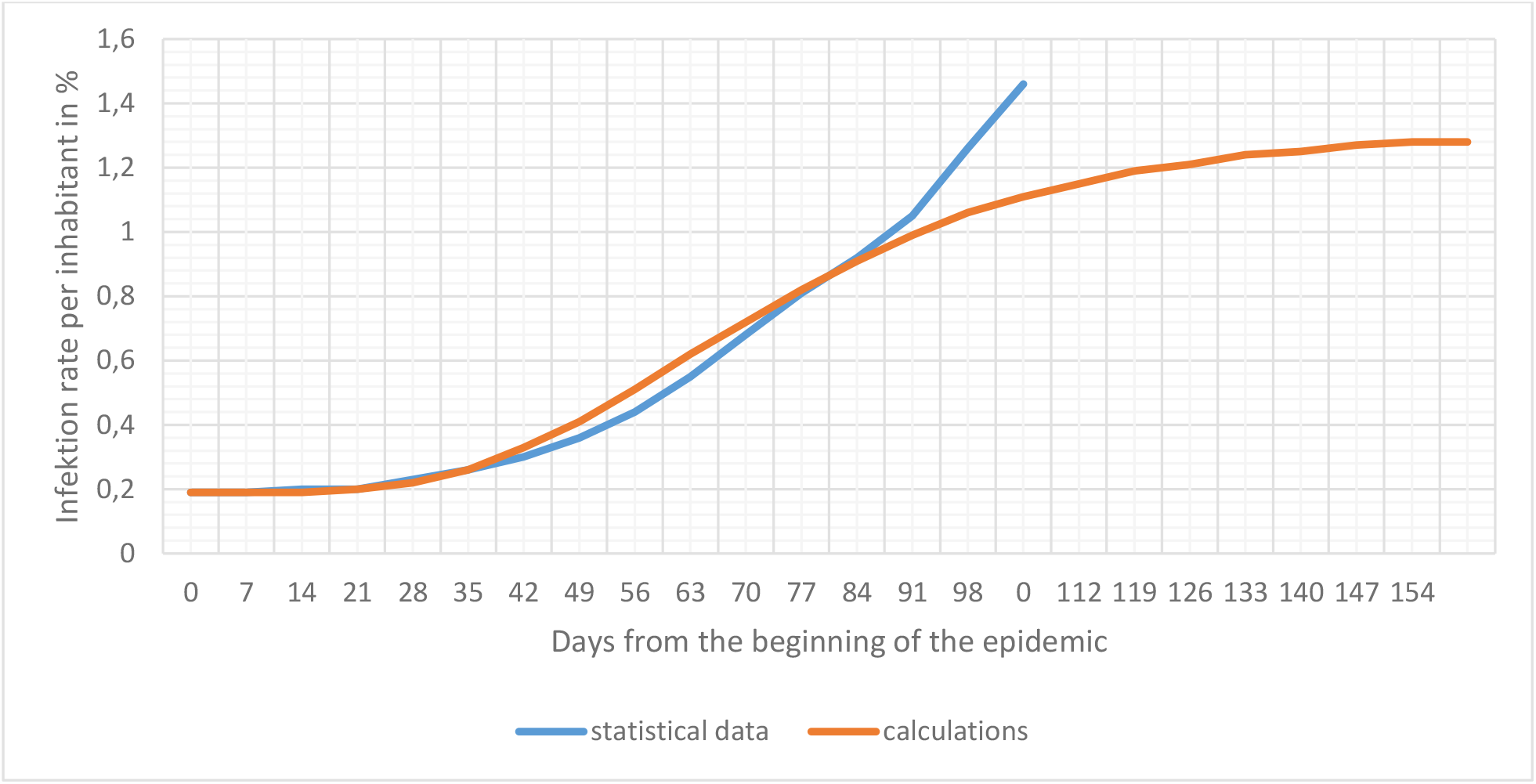
Relative growth of the coronavirus epidemic for the Treptow-Köpenick district

For the Mitte district (Fig. 7) some excess of the statistical data over the calculated data is observed closer to the end of December, yet it is not so noticeable. For this region, the correlation coefficient between the data reaches the value r = 0.97.

**Fig.7:**
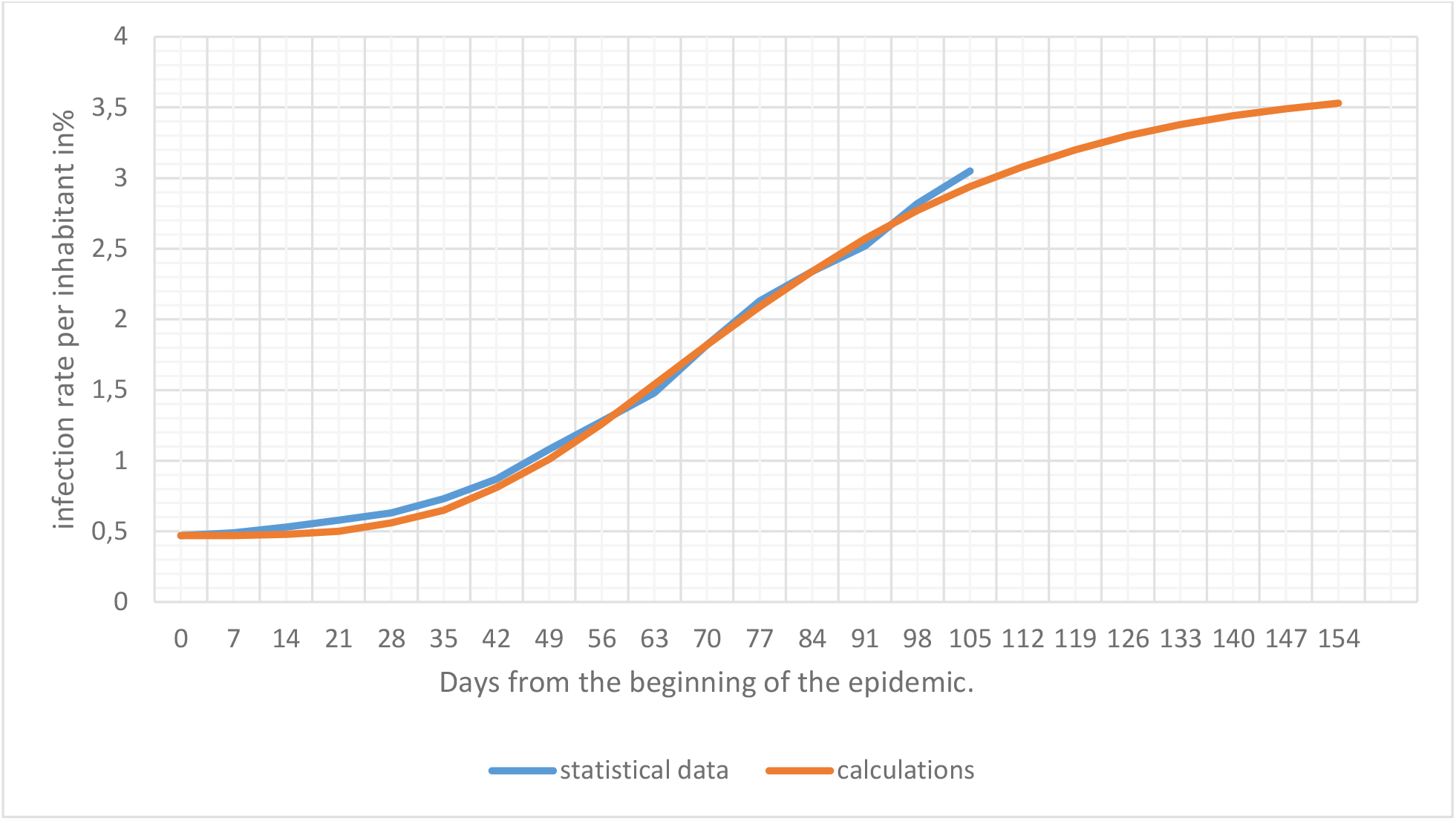
Relative growth of the coronavirus epidemic for the Mitte district

Thus, the data presented in Fig. 2 to Fig. 7 show that the results of calculations carried out according to dependencies (4) and (5) are in good agreement with the statistical data both for individual areas of Berlin, for the city in general and even for Germany. The only coefficient which allows getting such good agreement is λ. This coefficient varies in the range from 0.034 to 0.038 1/day.

Table 1 summarizes the data for all computed settlements.

**Table 1:**
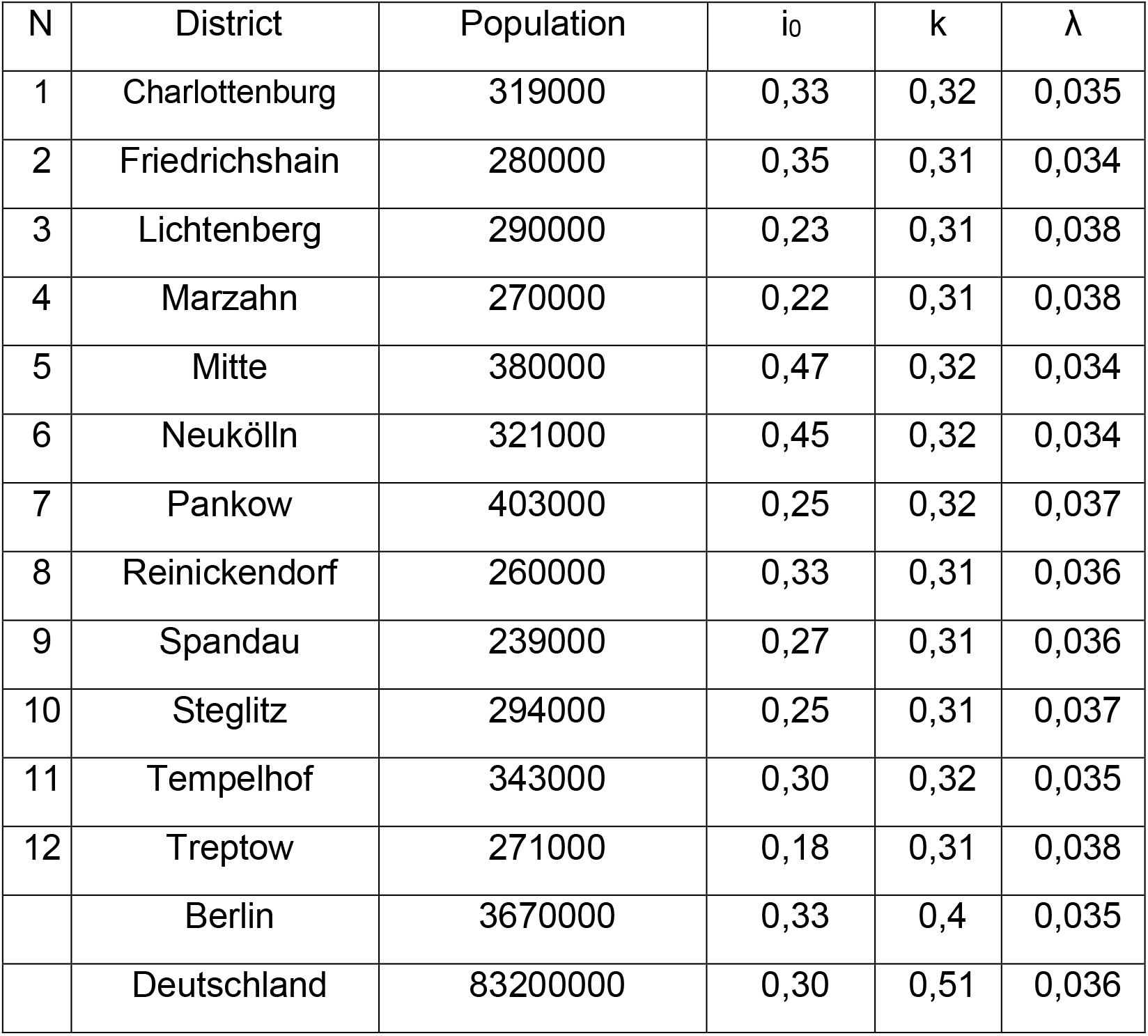
Summary of the data for all computed settlements

The accuracy of the calculations could have been increased by taking three significant figures for the model coefficients instead of two, but this was not necessary at this stage of the work.

The most important thing is to establish the relationship between the coefficient λ and socio-demographic factors that can influence the intensity of the epidemic spread.

Considering that one of the most likely places of transmission is apartments, where people communicate most intensively, we can assume that the coefficient λ should depend on the area per person, i.e. on the density of inhabitants in the apartments. For the Neukölln district this specific living area is 35.9 m^2^/person, but in Charlottenburg it is higher with 46.2 m^2^/person. Figure 8 shows a graph of the dependence of λ on this factor.

**Fig.8:**
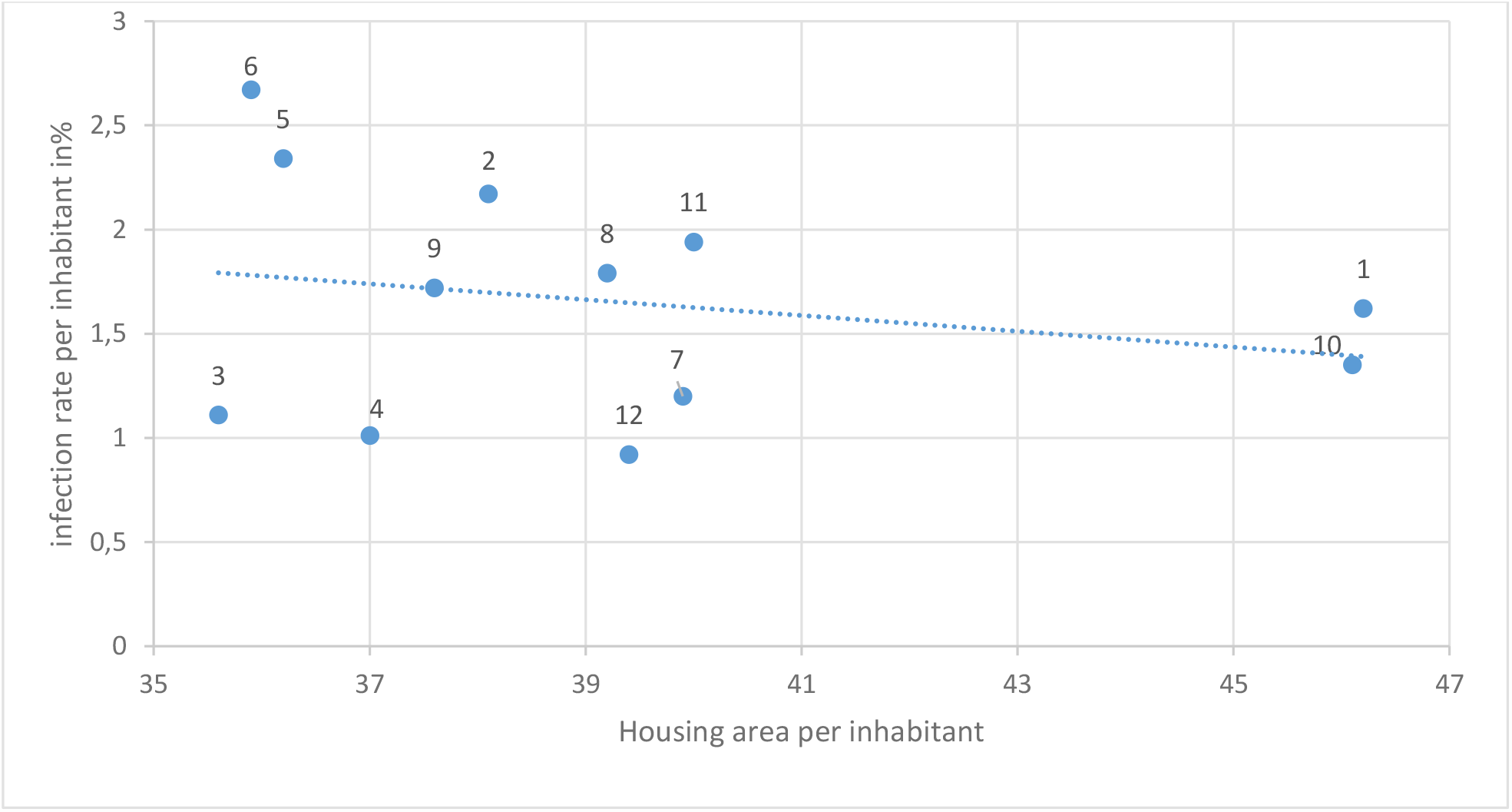
Dependence of the coefficient λ on the size of living space per person

Considering that one of the most likely places of transmission is apartments, where people communicate most intensively, we can assume that the coefficient λ should depend on the area per person, i.e. on the density of inhabitants in the apartments. For the Neukölln district this specific living area is 35.9 m^2^/person, but in Charlottenburg it is higher with 46.2 m^2^/person. Figure 8 shows a graph of the dependence of λ on this factor.

Contrary to expectations, the correlation between these parameters turned out to be extremely weak, the correlation coefficient being about r = 0.22.

The correlation coefficient was somewhat higher for the dependence of this coefficient on the value of total population density, that is, on the ratio of the total area of each of the districts to the number of inhabitants living in it. But even for these parameters the correlation coefficient turned out to be less than 0.7.

The graph of dependence of epidemic growth intensity on population density was presented in our previous work, where it was shown that the correlation coefficient of such dependence is equal to 0.67.

However paradoxical it may seem, it was found that the highest level of correlation was obtained when analysing the dependence of the number of infected people for each of the 12 districts in Berlin on the relative size of the population from the member countries of the Organization of the Islamic Conference (OIC) (Turkey, Arab States, some African countries) (r =0.93). In Figure 9 the dependence of the coefficient λ on this parameter is presented.

**Fig.9:**
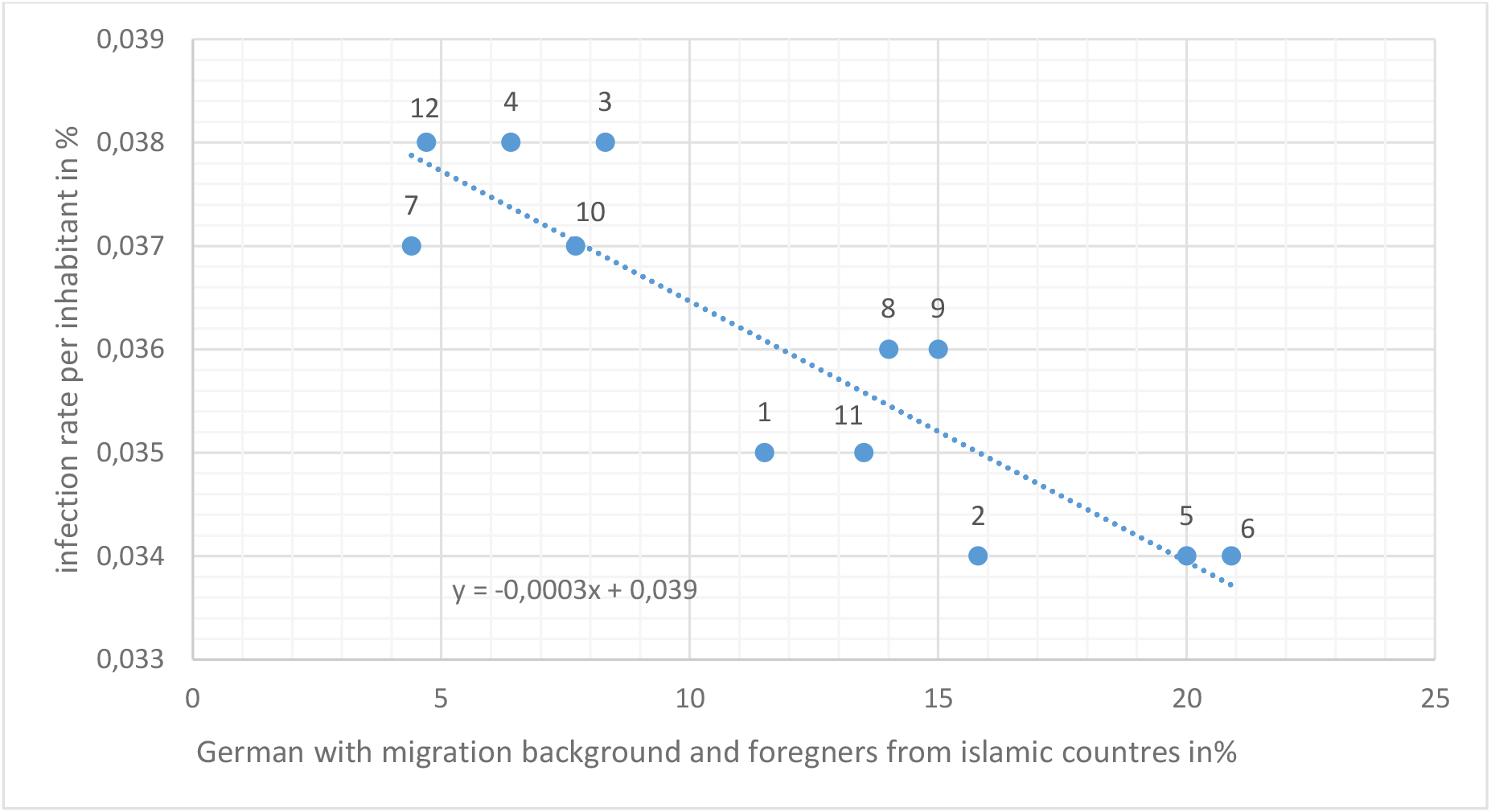
Dependence of λ on foreigners from countries OIC

The correlation coefficient between the parameters in this graph is r= - 0.89. At the same time, if the coefficient λ is taken to three significant digits, the correlation coefficient increases and becomes even higher. Such a high level of correlation between the parameters shown in Fig.9 raises questions. Apparently, foreigners from member countries of Islamic countries have closer family and friendship ties, which can contribute to more intensive transmission of the virus. It would be more correct to relate the intensity of transmission to certain psychological characteristics according to the theory on the behavioural immune system (BIS) [7]. However, the study of these problems is at the initial stage and special comprehensive psychological studies are needed to solve them. In order to study the influence of the main factors determining the rate of epidemic growth, hence having the maximum effect on the value of the λ coefficient, it is necessary to perform additional studies on the peculiarities of infratexture and socio-demographic characteristics for each of Berlin’s districts. At the same time, it is necessary to take into account the large variability of these factors within most districts, in which regions with a homogeneous structure can be additionally distinguished. In particular, one of these factors may be differences in the age distribution of the population in different districts. As was already established in our previous work, the maximum percentage of infected people are young people between the ages of 20 and 29 who are the most socially active. The disease rate in this age group reaches 1.5%. School-age children and people in the 30 to 50 age group are slightly weaker, slightly more than 1%. This conclusion was made based on the study of the epidemic in its initial phase. New data obtained for a longer period of the epidemic are shown in Fig. 10.

**Fig.10:**
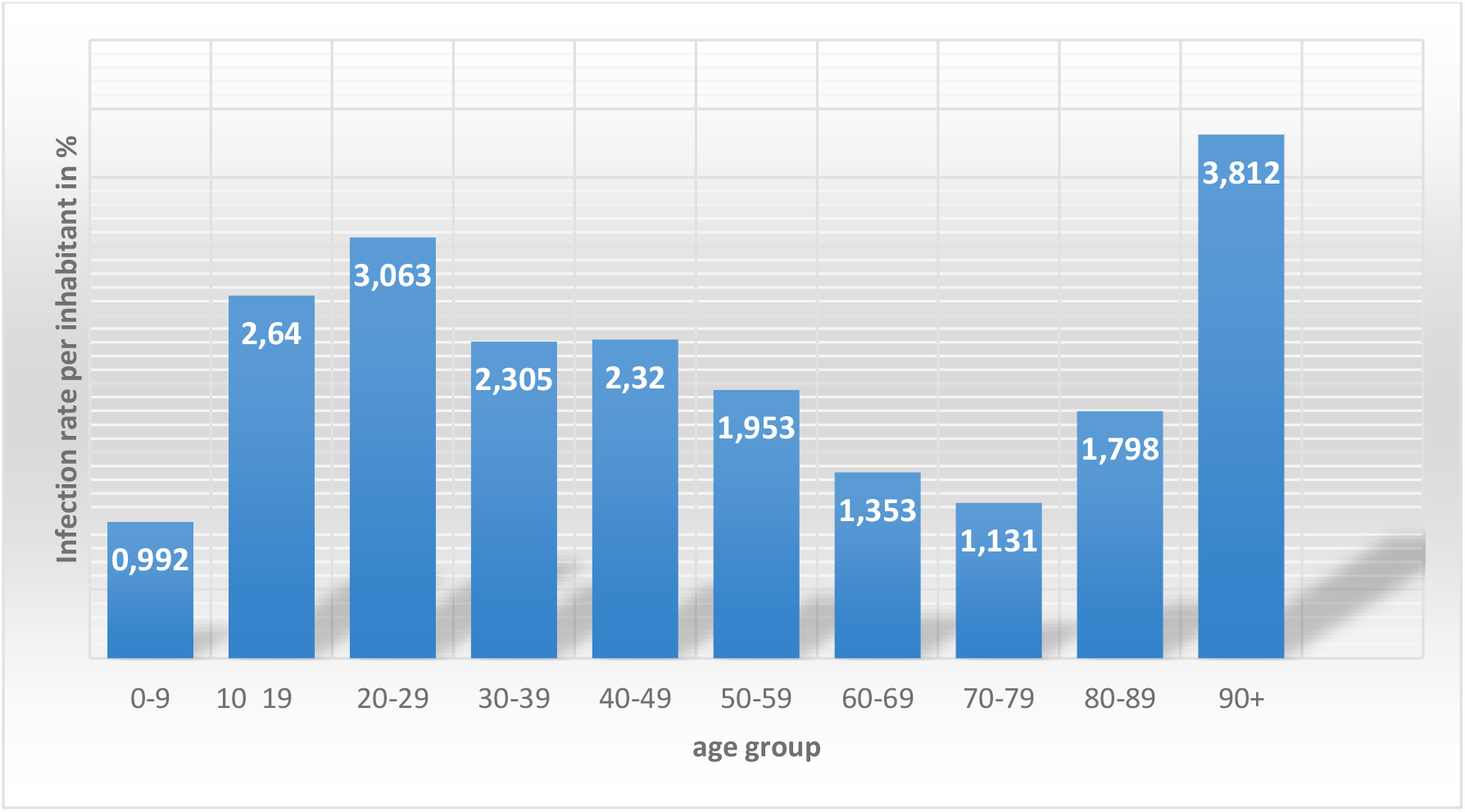
Relative number of infected people according to their age

At this stage of the epidemic the differences between the relative numbers of infected persons from 10 to 60 years old have slightly decreased. A more detailed analysis shows that more than 50% of all infected people (over 41,000) belong to the age group of 20 to 50 years old. The maximum percentage of those infected (3.36%) belongs to the most active age group of 20 to 24 years old. As for the elderly people 90+, although the relative number of those infected is quite high, the absolute number of people of this age group for Berlin is just over 1,000 people. In this regard, some doubts can be raised about the advisability of first vaccinating the elderly. Given that to date there is no reliable information about the possible effects of vaccination especially on immunocompromised individuals, it may be more appropriate to vaccinate the 18-30 age group in order to minimize the speed of the epidemic. Naturally, this most active age group generally has stronger immunity, which reduces the possibility of negative effects associated with vaccination.

The possibility that a new strain of the virus could emerge with a higher intensity of spread, which is of great importance for the further development of the epidemic, cannot be considered with a sufficient degree of certainty at the time of writing. Only speculations can be made at this point in the spread of the infection, since virologists have not yet discovered, either in Berlin or in Germany as a whole, a new virus strain which would differ considerably in its behaviour from the known strain. However, as noted above, the nature of the epidemic growth curves indirectly points to the possibility that such a strain of the virus could lead to a third wave of the epidemic.

In this case, the key question is whether the two characteristic strains of the virus can coexist simultaneously for a long time. It is possible that the more active strain displaces the “weaker” variant of the coronavirus. Unfortunately, virologists are not able to give an unambiguous answer to this question. If we assume that we currently have in Berlin an intermediate state of epidemic development, where a new mass strain of the virus emerges and begins to spread against the background of the existing epidemic, then an analysis of the intensity of infection growth can be performed using the ratio (5), by adding an additional summand to it:

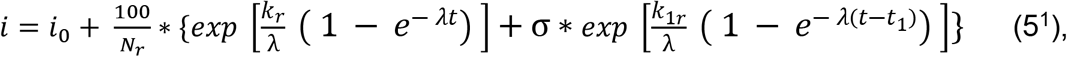

In which:

*K*_1*r*_ - Transmission rate coefficient of the new virus strain and the time of the epidemic wave associated with the new coronavirus strain

t_1_ - time of the beginning of the epidemic wave associated with the new coronavirus strain.

σ - Heaviside step function. σ = 1 when t ≥ *t* _1_ and σ = 0 when t < *t*_1_.

A preliminary calculation for different districts of Berlin according to ratio (5^1^), shows that the use of the proposed methodology will make it possible to successfully forecast the spread of the epidemic when two strains of the virus are exposed at the same time.

Fig. 11 shows the results of calculations for the Treptow-Köpenick district. As noted above, the maximum discrepancy between the calculated and statistical data was exactly this district.

**Fig.11:**
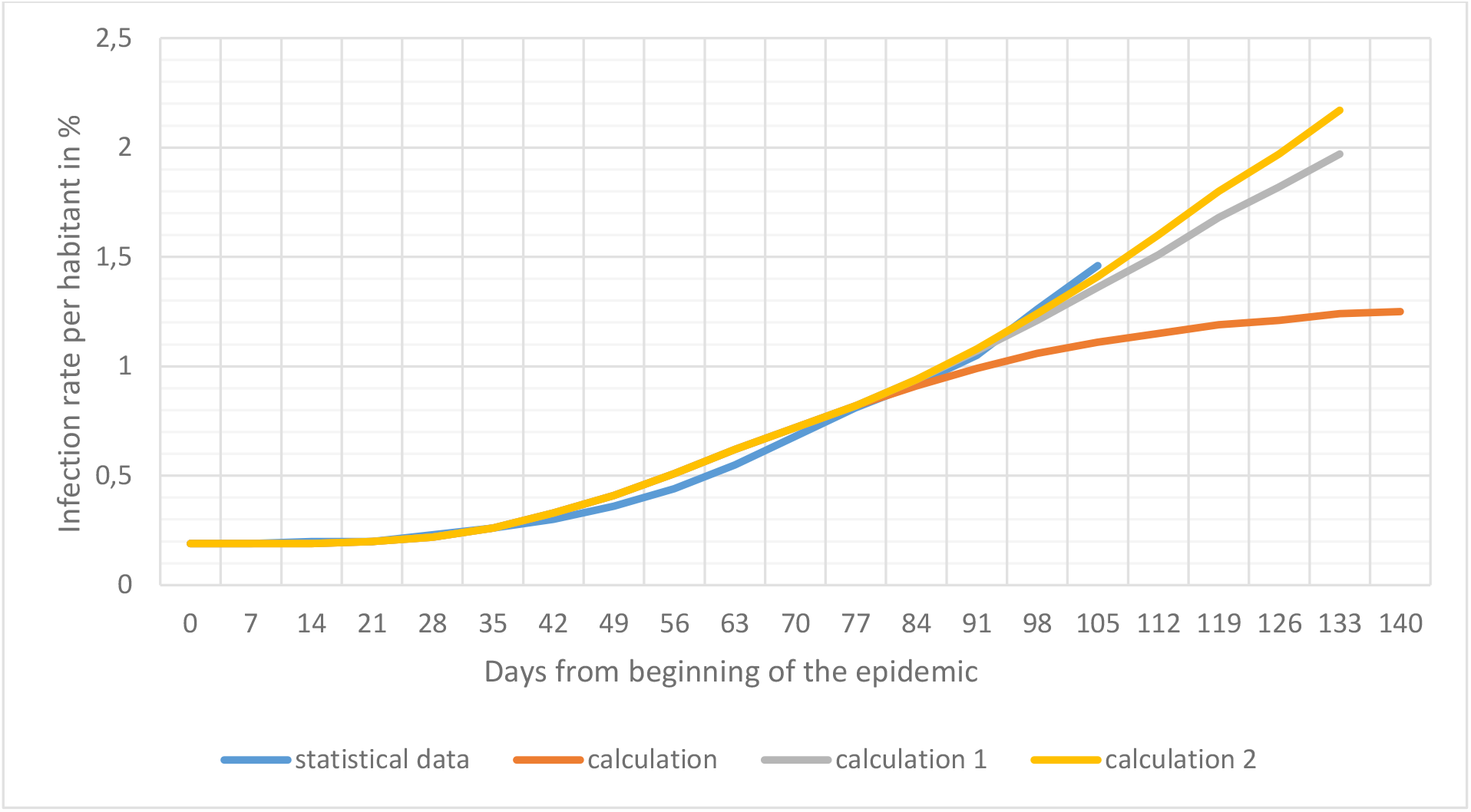
Relative growth of the two-strain coronavirus epidemic for the Treptow-Köpenick district

In addition to what was described above, calculation 1 was also performed for the condition that the new virus strain spreads with the same activity as the previous strain (i.e., *k*_1*r*_= 0.31) and calculation 2 for the assumption that the second virus strain is more transmissible (*k*_1*r*_= 0.32). The calculated data for the second assumption agree very well with the statistical data (r = 0.995).

It should be kept in mind that as time increases, the role of the first summand in the ratio (5^1^) decreases compared to the second, which takes into account the influence of the second strain however, a crucial question arises concerning the possibility of subsequent epidemic waves due to virus mutation. The virus, as studies by virologists show, mutates continuously. But only after a certain large number of mutations it acquires properties that significantly distinguish it from the previous strain. The time for the emergence of such, so to speak, new strain decreases as the intensity of epidemic growth increases. In other words, we can assume that the higher the derivative of the number of infected individuals over time, the faster we can expect the emergence of a new virus. However, this assumption would be true for a closed system. In reality, the emergence of a new strain of the virus is more likely due to its introduction from outside as a result of the continuing movement of people between different countries.

The proposed ratios do not assume the effect on the spread of the virus of mass vaccination of the population. However, in the case of such vaccination, the proposed calculation model can be easily modified to control its effect on the development of the epidemic. This is the undoubted advantage of analytical solutions compared to cumbersome numerical calculations.

In addition to the conclusions drawn in the preceding part of this paper [1], the following conclusions can be added:

1. The proposed model adequately describes the development of the coronavirus epidemic with insufficient adherence to quarantine and social distancing. The transition from the absolute number of infected persons to their relative number per inhabitant of a settlement makes it possible to obtain universal calculation ratios.
2. When performing the calculations, the choice of the date of the beginning of the epidemic is of great importance. Recommendations on how to determine the date of the beginning of the epidemic based on the analysis of statistical data on the spread of the epidemic are given.
3. The transmission rate coefficient k included in the calculated prognostic relation depends on the number of the examined population centre and the type of virus strain. A simple ratio for calculating this coefficient depending on the population size is proposed.
4. The control calculations performed, in which only a single empirical coefficient was used, showed high accuracy. The calculated curves for Germany, Berlin, and its districts agree well with the corresponding statistical data. The correlation coefficients between the corresponding curves reach values of 0.93 to 0.97.
5. The further development of the model should thus go in the direction of identifying causal links between the intensity of the epidemic and the main factors affecting this process. Some of these factors are related to the characteristics of the population’s behaviour and the infrastructure of cities.
6. The increase in the incidence in areas with a large percentage of the population rooted in Islamic countries is one of the main factors determining the development of the epidemic in Berlin. The analysis of the connection between this factor and the only empirical coefficient used in the calculations allowed us to establish that the correlation coefficient between them is equal to - 0.89. In order to explain and clarify this conclusion it is necessary to carry out further special socio-demographic research.
7. The age composition of the population also affects the speed at which the epidemic spreads. The maximum relative infection of the virus is observed in young people between the ages of 20 and 30.
8. Based on the comparison of statistical data with the results of the calculation, it is assumed that a new strain of coronavirus may have appeared in Berlin and Germany and that a new, third wave of the epidemic may have occurred.
9. The ratio for predicting the epidemic spread under conditions of simultaneous existence of both strains of coronavirus is given. Preliminary calculations show that, apparently, the spread rate of the new virus strain is somewhat higher than that of the previous one. Further improvement of the methodology of such calculations should be made on the basis of special virological studies.
10. The choice of a vaccination strategy for the population is questioned. It is suggested that vaccination of the most active age group will minimize the spread of infection, including among the elderly, while minimizing the risk of adverse side effects. Multivariate forecast on the basis of the proposed dependencies will allow to give recommendations on the most rational strategy of vaccination of the population.
11. The simplicity of the proposed prediction methodology and high accuracy of the results allow us to recommend it as a tool for operational analysis of various activities aimed at combating the spread of the COVID-19 epidemic.

## Data Availability

[1] Below, D., Mairanowski, J., & Mairanowski, F. (2020). Checking the calculation
model for the coronavirus epidemic in Berlin. The first steps towards predicting the
spread of the epidemic. medRxiv.
[2] Below, D., & Mairanowski, F. (2020). Prediction of the coronavirus epidemic
prevalence in quarantine conditions based on an approximate calculation
model. medRxiv.
[3] Der Regierende Buergermeister von Berlin, Senatskanzlei Informationen zum
Coronavirus (Covid-19). https://www.berlin.de/corona/lagebericht.
[4] European Centre for Disease Prevention and Control, Stockholm (2020). Rapid
increase of a SARS-CoV-2 variant with multiple spike protein mutations observed in
the United Kingdom, 20 December 2020.
[5] Amt fuer Statistik Berlin-Brandenburg (2020). Statistischer Bericht A I 16, hj 2/ 19
Einwohnerinnen und Einwohner im Land Berlin am 31. Dezember 2019. Potsdam.
[6] COVID-19 Dashboard by the CSSE at Johns Hopkins University (access on 28th
Dec. 2020)
https://www.arcgis.com/apps/opsdashboard/index.html#/bda7594740fd40299423467b48e9ecf6.
[7] Shook, N. J., Sevi, B., Lee, J., Oosterhoff, B., & Fitzgerald, H. N. (2020). Disease
avoidance in the time of COVID-19: The behavioral immune system is associated
with concern and preventative health behaviors. PloS one, 15(8), e0238015.

## References

[1] Below, D., Mairanowski, J., & Mairanowski, F. (2020). Checking the calculation model for the coronavirus epidemic in Berlin. The first steps towards predicting the spread of the epidemic. medRxiv.

[2] Below, D., & Mairanowski, F. (2020). Prediction of the coronavirus epidemic prevalence in quarantine conditions based on an approximate calculation model. medRxiv.

[3] Der Regierende Bürgermeister von Berlin – Senatskanzlei Informationen zum Coronavirus (Covid-19). https://www.berlin.de/corona/lagebericht.

[4] European Centre for Disease Prevention and Control, Stockholm (2020). Rapid increase of a SARS-CoV-2 variant with multiple spike protein mutations observed in the United Kingdom – 20 December 2020.

[5] Amt für Statistik Berlin-Brandenburg (2020). Statistischer Bericht A I 16 – hj 2/ 19 Einwohnerinnen und Einwohner im Land Berlin am 31. Dezember 2019. Potsdam.

[6] COVID-19 Dashboard by the CSSE at Johns Hopkins University (access on 28th Dec. 2020) https://www.arcgis.com/apps/opsdashboard/index.html#/bda7594740fd40299423467b48e9ecf6.

[7] Shook, N. J., Sevi, B., Lee, J., Oosterhoff, B., & Fitzgerald, H. N. (2020). Disease avoidance in the time of COVID-19: The behavioral immune system is associated with concern and preventative health behaviors. PloS one, 15(8), e0238015.

